# Using colorectal cancer screening evidence to stratify for personal patient risk among those with a family history of colorectal cancer: a 42-year cohort study

**DOI:** 10.64898/2026.06.04.26354891

**Authors:** Denis W King, Pamela E King, Megan Blanchard, Nickson Ning, Sebastian K King, Michael C Grimm, Tam Ha, Kathy Eagar

## Abstract

**Objective:** To determine if it is possible to assess individual patient risk of the development of colorectal cancer (CRC) in people in high-risk groups due to their family history.

**Design/Method:** Retrospective observational study of prospectively collected data from consecutive patients referred for a colonoscopy. 2,478 consecutive patients were referred to a single colorectal surgical practice in Sydney, Australia between 1977 and 2018 for a colonoscopy because of a family history of CRC. Of these, 1,963 have been followed for more than 10 years and are the subject of this paper. Histopathological findings categorised as normal (N), non-advanced adenoma (NAA) or advanced neoplasia (AN) with AN proven to be the precursor to CRC.

**Intervention:** Colonoscopic screening on the basis of contemporary practice to 2006 and subsequently according to Australian National Health and Medical Research Council guidelines.

**Results:** Participants with normal or low-risk findings in the first decade remain at lower risk of CRC for 30 years from the commencement of screening.

**Conclusion:** It is possible to stratify individual patients in a high relative risk cohort into those with high or low personal risk of CRC based on colonoscopic findings in the first 10 years of surveillance. Those with no AN in the first ten years have a lower 30-year risk of developing AN than the general community. This offers the possibility of structuring surveillance programs around individual risk rather than group risk, lessening the need for multiple surveillance colonoscopies in the majority of such patients and improving the cost effectiveness of CRC screening at the population level.

## INTRODUCTION

CRC is the third-most common cancer worldwide, and the second-most common cause of cancer death (1). The rate of CRC varies by country with a study of cancer registries finding that the age standardised incidence rate of early onset CRC is highest in Australia, USA, New Zealand, and South Korea. The rate of increase is faster among women than men in Australia (1). These variations suggest the need for intensified efforts to help facilitate early detection, particularly because screening has been shown to decrease death rates from CRC (2).

Family history has long been identified as a risk factor for CRC. However, other than in a small number of family cancer syndromes, where the specific genetic defect is known, there are no biochemical or genetic markers that may accurately predict which individuals within this high-risk group are likely to develop CRC.

This paper reports on a study in which CRC screening has been undertaken in routine clinical practice in accordance with Australian National Health and Medical Research Council (NHMRC) guidelines. These guidelines define three risk categories based on family history (3) as shown in Table 1.

**TABLE 1.** Colorectal cancer risk categories by family history, including degree of relationship, number of diagnoses in relatives and the ages of diagnoses, modified from jenkins et al. (3)

| Risk category – lifetime risk (to age 75 years if no screening) | Examples of asymptomatic people fitting into each category | Risk compared with the population average | Recommendations for screening |
| --- | --- | --- | --- |
| 1 — People at near average risk (5–10%) | • No FDR or SDR with CRC<br>• One FDR or one FDR and one SDR with CRC diagnosed at age $\geq 55$ years | • Risk is about 10% lower than the average risk<br>• Risk is about double the average risk, although most of that extra risk is expressed after the age of 60 years<br>• When the affected relative is second<br>degree (e.g., a grandparent, uncle or aunt), lifetime risk is only up to 1.5 times higher than average | • Biennial iFOBT from age 50–74 years.<br>• With one FDR $>55$ years consider biennial iFOBT from age 45 years |
| 2 — People at moderately increased risk (15–30%) | • One FDR with CRC diagnosed before the age of 55 years<br>• Two FDR or one FDR and at least two SDR diagnosed with CRC at any age | • Risk is about 3–6 times average risk.<br>• For the majority of people in this category, the risk of colorectal cancer is 3–4 times higher than average | • Biennial iFOBT from age 40–49 years, and colonoscopy every five years from age 50–74 |
| 3 — People at high risk (15–30%) | • At least three FDR diagnosed with CRC at any age<br>• At least three FDR or three SDR with CRC, with at least one relative diagnosed before age 55 years | • Risk is about 7–10 times average risk.<br>• For the majority of people in this category, the risk of CRC is 7 times higher than average | • Biennial iFOBT from age 35–44 years, and colonoscopy every five years from age 45–74 |

FDR – first degree relative
SDR – second degree relative
iFOBT - immunochemical faecal occult blood test

Most, if not all, CRC arises from adenomatous polyps that are initially benign, often for prolonged periods, and therefore amenable to detection and definitive treatment (4). The most common method for their detection and removal is colonoscopy.

Studies over more than 40 years have consistently found strong evidence in favour of endoscopic screening. These include the 1993 National Polyp Study (5), a 2001 population-based, state-wide study in Western Germany (6) and the Prostate, Lung, Colorectal and Ovarian (PLCO) Cancer Screening Trial (7).

The US National Polyp Study (NPS) was a randomised clinical trial which began accrual in 1980. In a review at 40 years, Winawer et al. (8) noted that the challenge at the beginning was to determine whether patients with a low risk of subsequent cancer could be identified for less-intensive surveillance. In that paper, the authors argued that repeated studies had validated risk stratification and expanded the low-risk surveillance intervals to five to ten years. They suggested that the relationship of continued surveillance to the findings at the first surveillance interval could reduce the burden of surveillance colonoscopy and provide more efficient use of resources for screening and diagnosis.

They also posed the question about how identification of advanced adenoma in a patient could be used clinically. They suggested that there was a need for long-term studies of varying surveillance intensity to better elucidate the surveillance required for low and high risk patients, such as the European Polyp Surveillance Study (EPOS) was one study which could inform the relationship of the advanced adenoma to CRC as an outcome measure and as a continued valid screening goal.

The debate about stratification has involved two key issues. The first is which clinical findings are most appropriate for risk stratification purposes (9, 10) (11). The second is the length of time for which any such stratification remains valid (6) (12, 13). This study addresses the latter issue in particular, with the observation period of this study matching the currently recommended lifetime period of surveillance.

A further issue is adherence to colonoscopy screening guidelines. The rate of adherence to recommendations is a constant problem with CRC screening of all types, with rates as low as 30% reported (14) but seldom more than 50% (15, 16). The evidence is that the more clearly a risk can be defined, the more likely the patient is to adhere to recommendations (17, 18). This suggests that a study which clarifies individual patient risk may improve adherence to recommendations.

## MATERIALS AND METHODS

This is a longitudinal observational study in a single colorectal surgical practice in Sydney, Australia. Participants were referred to the practice between 1977 and 2018 for a colonoscopy either because they had a family history of CRC (FHCC), a personal history of adenoma, or a personal history of CRC. This study only examines the FHCC cohort.

Screening for CRC is a research topic that, for both practical and ethical reasons, does not lend itself to a randomised controlled trial (5, 19, 20). Despite its acknowledged weaknesses, a longitudinal observational study seems an appropriate option.

Every patient with a FHCC was invited to participate as part of their routine treatment with no financial incentive or penalty. Patients who had pelvic radiotherapy previously or had total ulcerative colitis were excluded due to the increased CRC risk associated with those conditions. This ensured that there was no selection bias. The consent process was part of routine clinical treatment for all subjects and included specific consent for the collection of all relevant information for the purposes of research and publication.

The history of colorectal disease and interventions was obtained directly from the participant at the initial consultation. All members of the practice who had direct contact with patients about their treatment were medical or nursing health professionals experienced in the management of colorectal disorders.

Recruitment of consecutive participants started in 1977 and finished in 2018. Participants were followed up until contact was lost, or until 31 December 2024. All participants were treated at some time during their period of surveillance by the principal author and all participants who were followed up for a minimum of ten years were included in the study. The vast majority of the procedures were performed by the author personally or by Accredited Trainees of The Royal Australasian College of Surgeons or Colorectal Surgical Society of Australia and New Zealand working under direct supervision. The index colonoscopy for each patient was the first performed at or subsequent to the recommended age for the commencement of surveillance.

All polyps estimated to be greater than 10 mm, based on visual comparison at colonoscopy to the 7 mm spread of hot biopsy forceps, were resected completely and sent for histological examination. For reporting purposes size was based on the findings in the fixed specimen where available. For those that were clearly less than 10 mm in diameter, the majority were treated using biopsy forceps, and after the initial few years, hot biopsy forceps.

The histopathological findings were categorised as normal (N), non-advanced adenoma (NAA), and advanced neoplasia (AN), as shown in Table 2. The critical pathology parameter is the presence or otherwise of adenomatous polyps. Those cases in which the distinction was in doubt were reviewed by a meeting of the practice surgeons and accredited surgical trainees, and the pathology providers, usually with 8-10 surgeons and 3-4 pathologists. A decision about the final pathology report was made after discussion and by consensus.

**TABLE 2.**
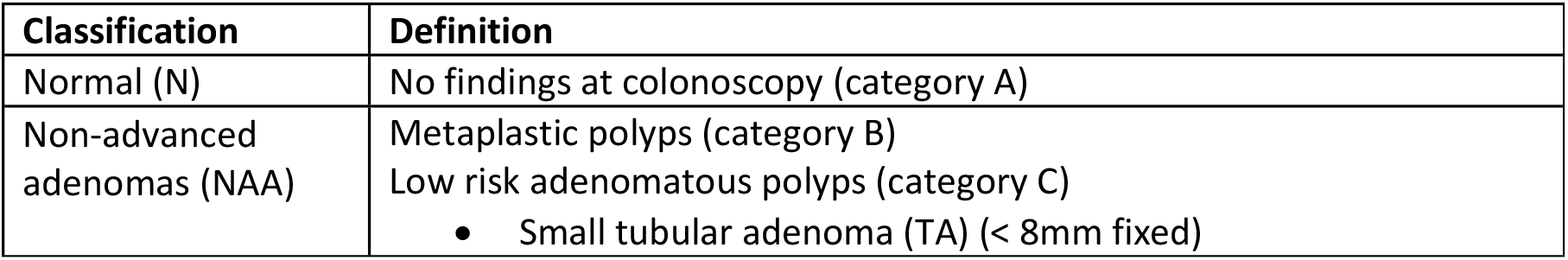
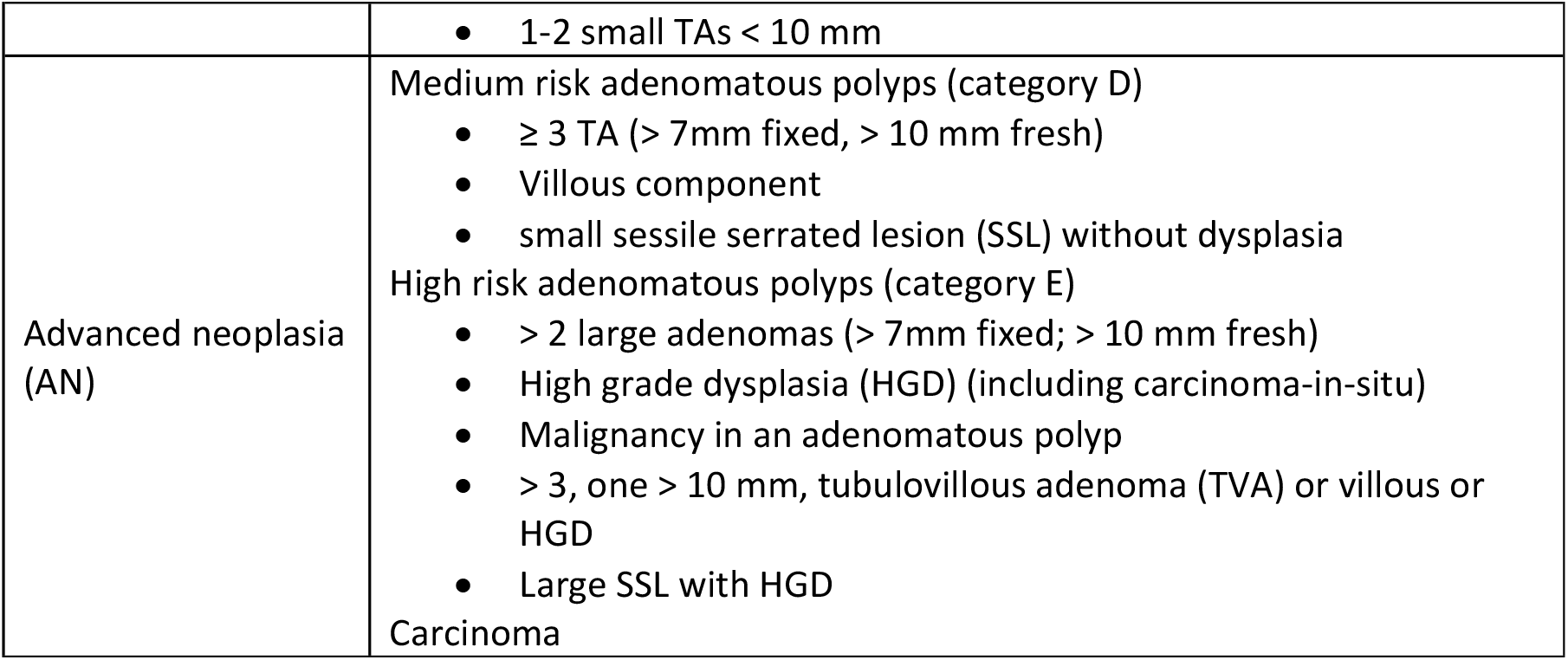
Categorisation of findings at colonoscopy, modified from lieberman et al. (21)

The procedure involved, colonoscopy with standardised preparation, was uniform for all participants, irrespective of the specific indication, or of the outcome of the investigations. No stratification was used during the study and recommendations for surveillance intervals were based on entry criteria and ongoing findings. A comprehensive follow-up and recall system was applied equally to all participants under surveillance and was managed according to fixed protocols by practice staff, independently of the principal author (22).

In the first half of the analysis survival curves were calculated using the life table (actuarial) estimate of the distribution, with 95% confidence intervals (CI). Life table estimates were chosen over Kaplan-Meier as the actual date of the development of a polyp was not known, but rather the date of the colonoscopy at which it was observed. Survival analysis was performed using the year post-index colonoscopy as the ‘time-to-event’ variable. Data were analysed using SAS version 9.4 and extracted into Microsoft Excel (Microsoft, Seattle, USA) for graphing.

In the second half of the analysis, survival curves and pointwise tests were calculated at monthly intervals using Greenwood’s formula. For the cumulative number, size, and area metrics, Welch’s t-tests were used. De-identified information from the database was exported into a Microsoft Excel spreadsheet for analysis and coded by high-risk category. Survival curves were compared using the log-rank test and the null hypothesis rejected (P < 0.0001).

## Ethics approval

The Human Research Ethics Committee (HREC) of the University of Wollongong, Australia approved the project in 2020 (approval 2020/386), after the completion of recruitment, with ratification by the University of New South Wales HREC in April 2023.

## RESULTS

A total of 5,056 participants with a perceived increased risk of CRC due to a family history of CRC (FHCC), a personal history of adenoma, or CRC were recruited between 1977 and 2018. Of that group, 2,478 had a family history of CRC. Of this FHCC cohort, 1,963 participants were followed for more than 10 years with an average length of follow-up of 25.22 years and a median of 24.97 years (IQR 17.64–31.62). The FHCC cohort underwent a total of 8,761 colonoscopies with six perforations and no deaths. The completion rate to the caecum was 98.73% (incomplete 111).

The demographic characteristics of the participants match the catchment population from which the patients were largely drawn, and Australia as a whole, other than for a slight over-representation of females in the study population. The median age of 59 years is considerably older than the overall population, because surveillance is usually not recommended until the age of 50 years. More detail on the study population is provided in the supporting information file.

Figure 1 shows results from two groups based on the findings in the first 10 years. One group are those with normal or NAA. The other is those with a confirmed finding of an advanced neoplasm in the first 10 years. Each patient developing an AN is entered once only. The incidence rates for each group are the number of participants who developed any new AN in the observation period as a proportion of the number of participants remaining in that group each year since the index colonoscopy.

**FIGURE 1.**
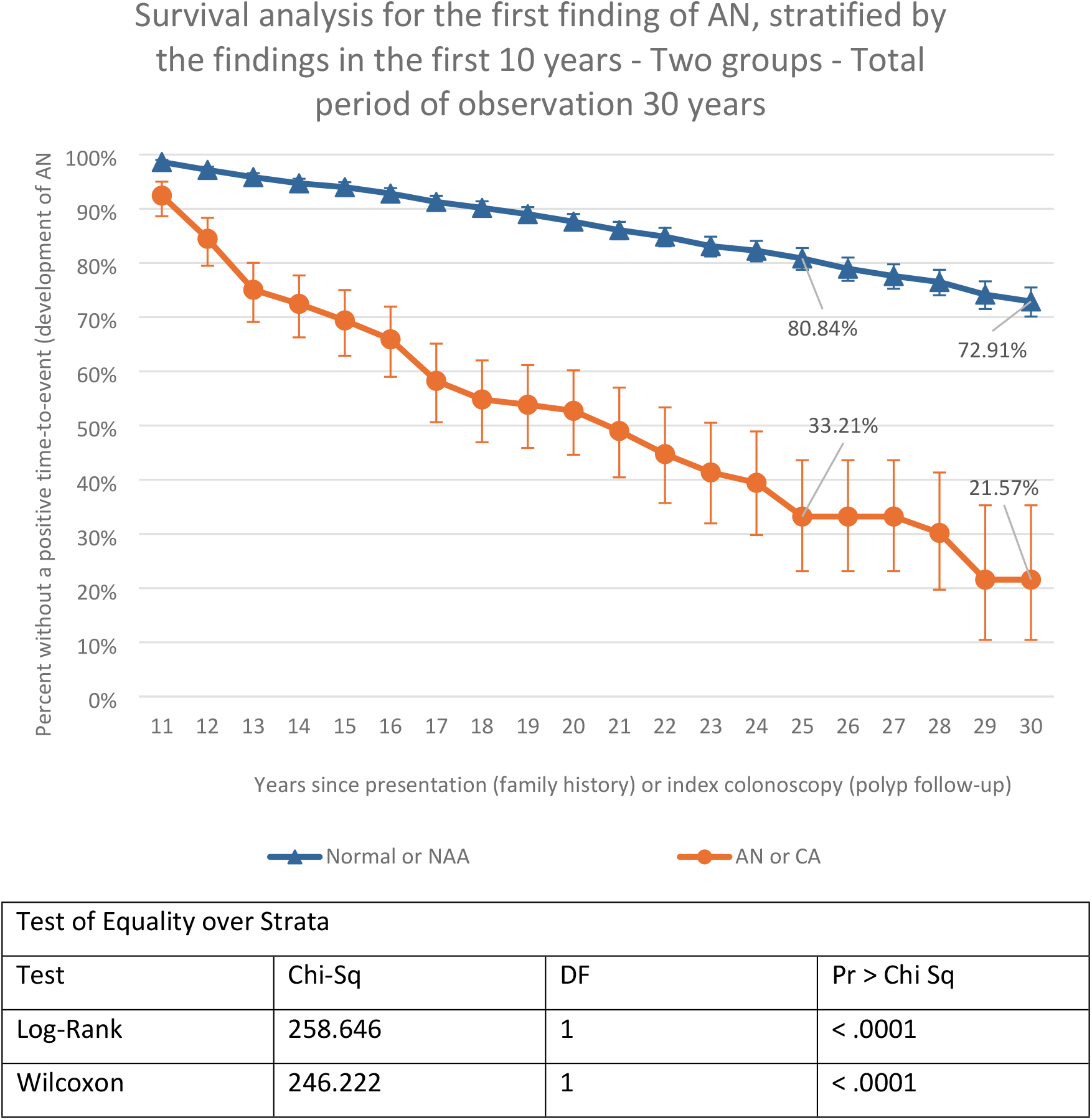
Survival analysis for the first finding of an, stratified by the findings in the first 10 years.

Figure 2 shows the cumulative proportion of participants who developed AN in the three groups: those with AN in the first 10 years, those with NAA in the first 10 years, and those with normal colonoscopies in the first 10 years. The graphs represent the cumulative total of the development of AN during the study period. The findings in the first 10 years are significantly different (p < 0.0001) between the AN and the other two groups, and the disparity progressively increases for 30 years. For the purposes of assessment of development of CRC, the AN in the first 10 years are included in the overall results. This is based on the long lead time from the development of AN to the transition to carcinoma. The cumulative incidence of AN in the high-risk group at stratification was nearly four times as high as those with NAA and twelve times as high as those with no neoplasia in the first 10 years.

**FIGURE 2.**
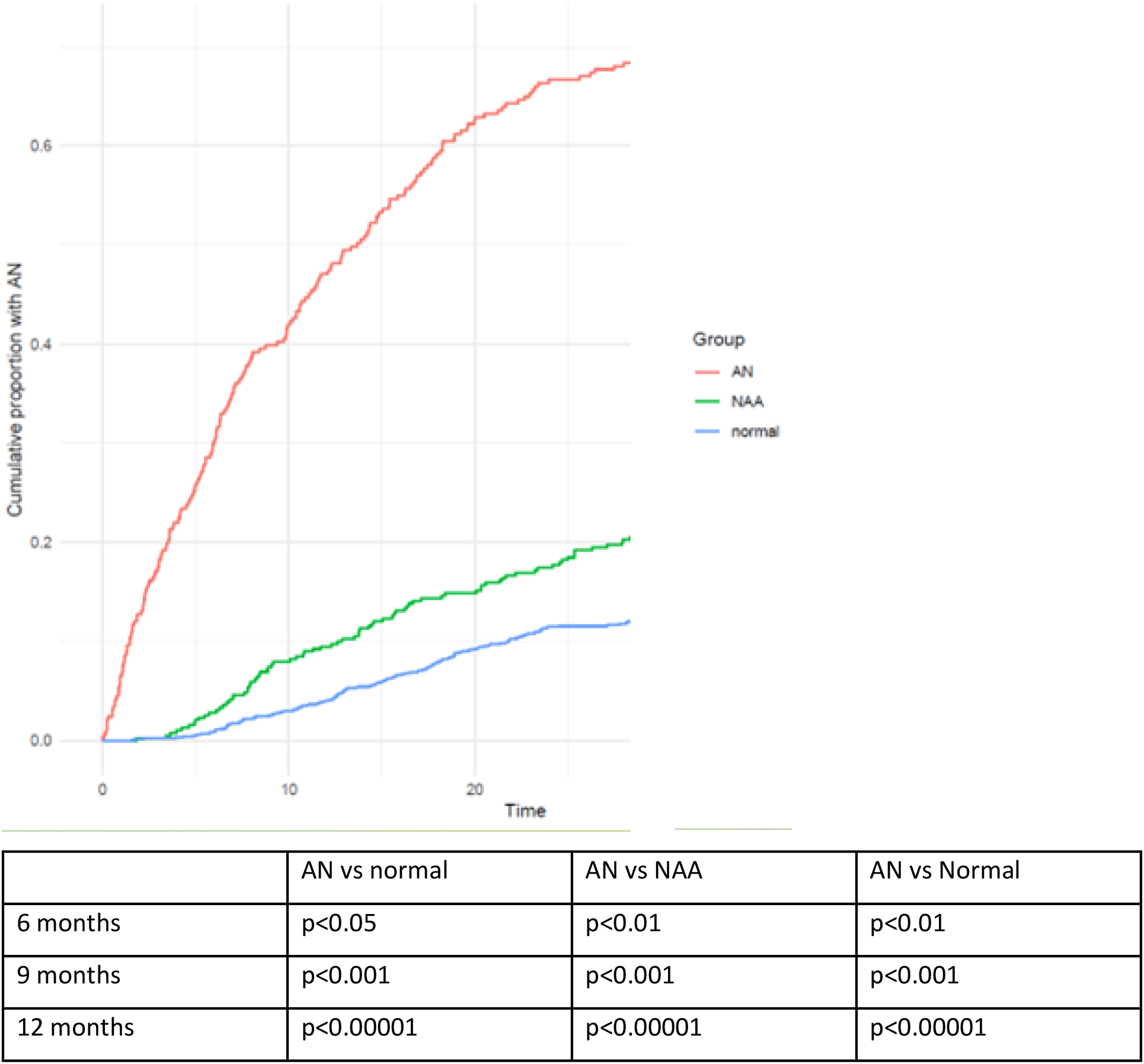
Cumulative proportion of patients with an.

## DISCUSSION

Patients with a FHCC with normal or low-risk findings in the first decade, that is no AN, remain at lower risk for 30 years from the commencement of screening. The development of AN in the first 10 years indicates the likely continuing development of AN and presumably therefore increased risk of carcinoma for the full period of currently recommended surveillance protocols (5, 23).

These findings confirm that it is possible to stratify individual patients who are in a high relative risk cohort into those with high or low personal risk of CRC based on colonoscopic findings in the first 10 years of surveillance. Those demonstrated to be at normal or low risk appear to have a lower risk of developing AN than the general community despite their family history (11, 24-27).

### Strengths and weaknesses of the study

A key strength of this study is that it reports the longest and largest study of an issue of singular importance in CRC surveillance. The outcome is a rational stratification method that is valid for the 30–year period of screening programs currently recommended by national bodies, including the Australian Cancer Council (28).

This study contributes to new knowledge by identifying those with a family history of CRC who are not at a high personal CRC risk. As well it means that patients may be informed of a personal risk rather than the more general group risk. At the patient level, this information can be used to reassure patients and subject them to CRC screening only if they are at significantly increased risk. Understanding individual patient risk, rather than group risk, improves adherence to recommendations (14, 29, 30).

At the system level, it means that patients with a genuine higher personal risk of CRC can be better targeted and effectively screened using less intrusive, community-based protocols. This has the potential to have a major impact on the cost-effectiveness of CRC screening with little or no attendant loss of clinical efficacy.

### Strengths and weaknesses in relation to other studies

The findings are consistent with previous studies such as those of Winawer et al. (5) and Zauber et al. (23) which indicated that AN is a marker of CRC potential. It differs from previous studies in that it has a much larger sample and a much longer observation period. For example, Wieszczy et al. (10) reported a mean follow-up of 7.1 years and a maximum follow-up of 14 years.

### Meaning of the study: possible explanations and implications for clinicians and policymakers

CRC is theoretically preventable because it has a known benign precursor suitable for detection and removal (15).

The outcome of this study is a stratification method to better predict which of those people with a FHCC will go on to develop CRC. It is valid for the currently recommended CRC surveillance period.

This is against the background of concerns about the number of potentially unnecessary colonoscopies performed, the risks to patients, and the load upon health systems of current surveillance protocols (31). Adoption of this approach in routine practice would improve the sensitivity and specificity of CRC screening after the initial ten-year period. This would allow resources to be targeted to those at actual risk. Those deemed to be at low risk would be spared the inconvenience and potential harm of colonoscopic screening, with no loss of its overall clinical efficiency.

In a paper entitled “*The National Polyp Study [NPS] at 40: Challenges Then and Now*”, Winawer et al. (8) documented challenges at the commencement of the NPS, those that have been resolved, and those that remain. Among other issues, they ask “Can surveillance intervals be further lengthened?

Can patients with low risk for cancer be identified for less intensive surveillance? How can the identification of an advanced adenoma…be used clinically?” This study addresses those questions.

This offers the possibility of structuring surveillance programs around individual risk rather than group risk, lessening the need for multiple surveillance colonoscopies in the majority of such patients. Studies have shown that, in relation to adherence to recommended guidelines, patients respond better to specific personal risk data then to generic group risk data, thus improving the clinical effectiveness and cost-effectiveness of screening programs (11, 18).

## Data Availability

1. The minimal data set [and accompanying code, where generated] is available at UNSW data repository via unsw.edu.au

## Statements and declarations

## Acknowledgements

We would like to thank Sydney Colorectal Associates, Sydney, Australia for supporting this study as part of routine clinical care, and for continuing to care for the patients following the principal author’s (DWK) retirement from practice (at the end of 2018).

We would also like to acknowledge the support received through an Australian Government Research Training Program Scholarship.

